# Social-economic drivers overwhelm climate in underlying the COVID-19 early growth rate

**DOI:** 10.1101/2021.09.10.21263383

**Authors:** Zhenghua Liu

**Affiliations:** School of Minerals Processing and Bioengineering, Central South University, Changsha 410006, China

## Abstract

Identifying the drivers underlying the spatial occurrence and spreading rate of COVID-19 can provide valuable information for their preventions and controls. Here, we examine how socio-economic and climate drivers affect the early growth rates of COVID-19 in China and the other countries, the former of which have consistently stricter quarantine during early epidemic and thus are used to enquire the influences of human interventions on trainsimissions. We find that the early growth rates of COVID-19 are higher in China than the other countries, which is consistent with previous reports. The global spread is mainly driven by the socio-economic factors such as GDP per capita, human movement and population density rather than climate. Among socio-economic factors, GDP per capita is most important showing negative relationships in China, while positive in the other countries. However, the predicability of early growth rates by socio-economic and climate variables is at least 1.6 times higher in China’s provinces than the other countries, which is further supported by metapopulation network model. These findings collectively indicate that the stochasticity of transimission processes decrease upon strict quarantine measures such as travel restrictions.

**Key findings:**

1. GDP per capita is most important in driving the spread of COVID-19, which shows negative relationships within China, while positive in the other countries.
2. Socio-economic and climate factors are key in driving the early growth rate of COVID-19, while the former is more important.
3. Socio-economic and climate features explain more variations of early growth rates in China due to the decreased stochasticity of transimission processes upon strict quarantine measures.

---

The coronavirus disease 2019 (i.e., COVID-19) has developed into a pandemic with an unprecedented speed since its outbreak in 2019 and exerts a great threat for public health at the global scale [1]. Geographical and climate factors are frequently reported to be important for the spread of COVID-19, such as latitude, temperature, humidity [2-5], and seasonality [5]. This is not surprising as climate is usually related the plague infection activity, such as SARS [6], MERS [7], Ebola [8], Zika [9] and influenza [10].

Socio-economic developments, however, could also profoundly influence epidemic transmission dynamic [10-12]. This is indicated by the historical co-occurance of increasing frequency of epidemics with recent human population (Fig. S1), which is largely because the socio-economic status relevant to rapid population growth [13], human movements [14, 15] and uncontrolled urbanisation [10] could provide conditions and environments conducive to vector proliferation and plague transmissions [16]. For instance, at regional scale, COVID-19 spread is promoted by city size in USA [17] and air pollutants of nitrogen dioxide [18] and PM2.5 [19], and is constained by control meausres, such as travel restrictions [20]. At global scale, COVID-19 cases could be seeded globally [15, 20, 21] via the well-developed traffic networks composed of railways, highways, waterways and airways across cities, provinces, states and countries. With the increasing movement of infected humans, virus spread and persistence are improved and the spatial dimensions of transmission are extended from small household foci to large community clusters [22, 23], thus leading to the recent expansions of COVID-19. Identifying socio-economic factors and their interactions with climate in underlying the worldwide COVID-19 spread will be helpful to understand its spatial patterns and provide useful information regarding its prevention, mitigation and control.

Here, we estimated and compared the early growth rates of COVID-19 between China and the other countries because the former could be regarded as a treatment group in respective of its stricter measures in social distanting and travel restictions especially during early epidemic. For each region at the province level of China or the state level of the other countries, we calculated a series of growth rates based on exponential model with extensible windows starting from the first confirmed case of COVID-19 and regarded the maximum value as early growth rate (Fig. S2, S3). The maximum growth rate is expected to locate on the inflection point of the slope in logistic growth curve during the exponential phase, where the growth of COVID-19 is free to control measures and natural resource limitations such as susceptible population, and thus is mostly closed to the theoretical growth charateristics of virus [24]. Different from the other counties, the growth rates in China were estimated at the province level, since strict quarantine was consistently applied across the country although there is a high spatial heterogeneity in climate and socio-economic status. Further, we explored the underlying drivers of COVID-19 early growth rates with the compiled 30 socio-economic and 16 climate factors (Table S1) based on 33 China’s provinces and 29 countries outside of China after. More details of growth rate estimate and environmental variables are shown in supplementary materials and methods.

As of 26^th^ Mar 2020 when the daily COVID-19 cases data were collected, there were more than 82,034 and 447,557 infected individuals confirmed in China and the other counties, respectively (Fig. S4a). The early growth rates in China were 0.495 ± 0.189 (mean ± SD), and significantly higher than the other countires (0.335 ± 0.167; two-side T-test, *P* < 0.001; Fig. S4b). This result agrees with the previous studies showing the higher basic reproduction number in China (3.38 ± 1.41) than the other countries (2.92 ± 1.02; Table S2) and the lower doubling time in China (2.16 days) than USA (2.46 days) and European countries (2.63 days) [25]. Outside of China, there was a significantly negative correlation between early growth rate and temperature or UV radiation (all *P* < 0.05; Table S3; Fig. S5), which is consistent with a significant geographic pattern of the increasing early growth rate towards high latitude (Pearson’s r = 0.383, *P* = 0.04). However, the above relationship between early growth rate and temperature or latitude was not observed in China (*P* > 0.05; Fig. S5; Table S4). Additionaly, it was worth noting that there were significantly unimodal relationships between growth rates and precipitations (*P* < 0.05; Fig. S5) across the other countries, but not in China. Such non-significant effects of climate variables in China are consistent with recent studies that the COVID-19 spread in China was independent on temperature and humidity [26, 27]. But it is contrasting with other reports showing that climate factors such as temperature and precipitation determine the spatial and temporal variations in the incidences and transimissions of most epidemics [28], such as malaria [16], cholera [29], and Ebola [30].

Compared to climate, socio-economic factors played more important roles in driving the spread of COVID-19 (Fig. 1, S4b, S6). Among them, GDP per capita was most important (Fig. 1a) and explained 19.5% and 8.9% of the varaitions of early growth rate in China and the other countries (Fig. 1b), respectively. In China, GDP per capita negatively correlated with early growth rate (Fig. S6; Table S4), while positively for the other counties (Fig. S6; Table S3). Such contrasting effects of GDP per capita may be resulted from the control measures such as travel restinctions, which disabled the human mobility and COVD-19 transmissions [20]. This is further supported by the commuting rate (Fig. S6), which showed no relationships with the growth rates in China, but significantly increased the rate in the other countries (r = 0.543, *P* = 0.002; Fig. S6; Table S3). This result indicates that in the regions with high GDP per capita, COVID-19 spread is promoted by attracting more immigrations [31] and increasing commuting rate (Fig S7), but COVID-19 transmission could be also effectively prevented by the high investments on human interventions such as travel restrictions and isolation of suspected cases, confimed cases and contacts.

**Figure 1.**
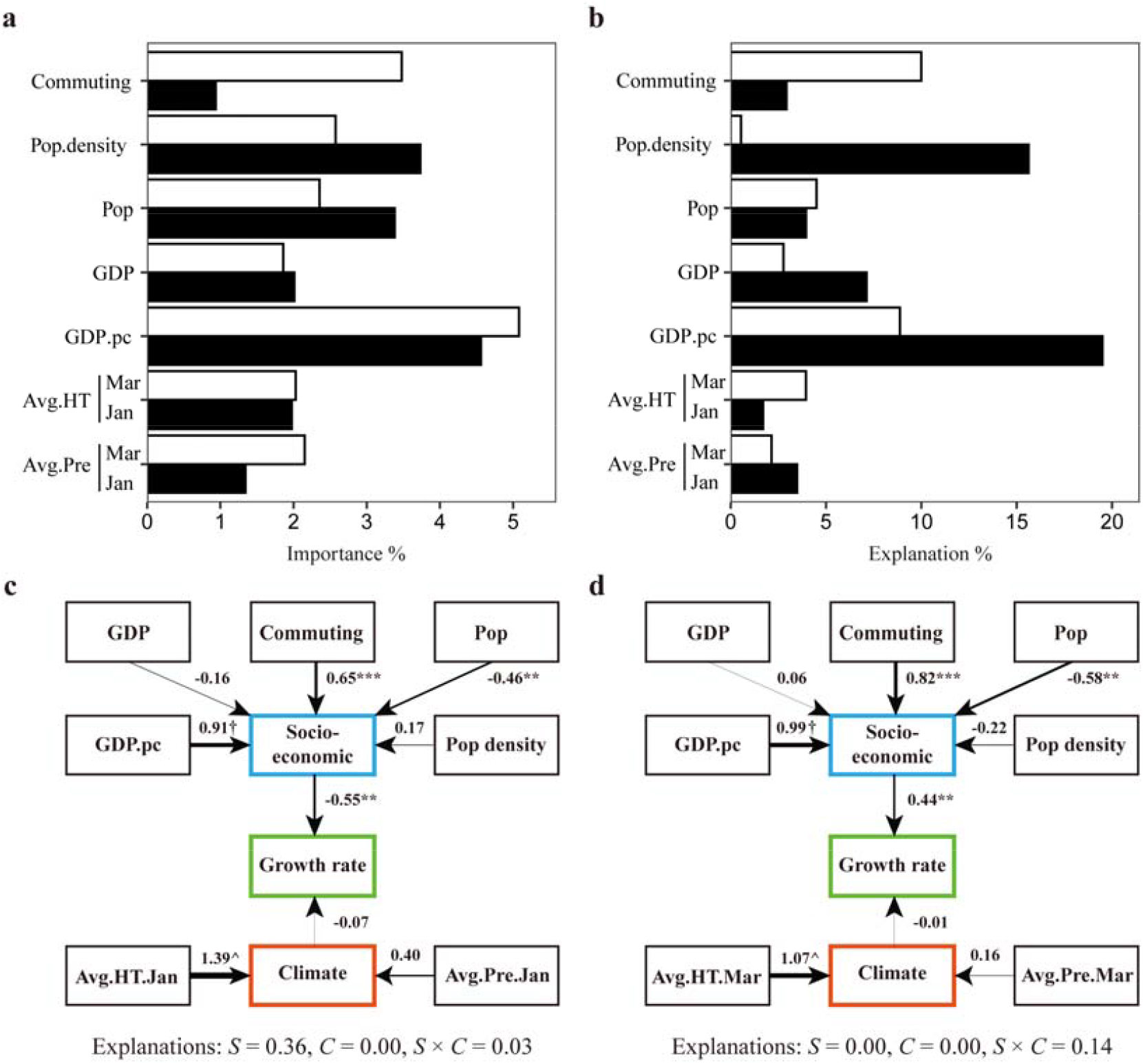
The roles of climate and socio-economic variables playing in the early growth rates of COVID-19. The relative importance of variables in early growth rate were estimated by random forest (**a**), while the individual explanations were calculated by redundancy analysis with the hierarchy algorithm (**b**). The total of variation explained by this six factors were 54.3% and 32.8% at the province level of China (black bars) and the state level of the other countries (white bars), respectively. In structural equation model (SEM) of early growth rate driven by socio-economic and climate factors at China’s province level (**c**) and state level (**d**). The blue and red windows represented the latent variables of socio-economic and climate, respectively. The explanations of socio-economic (*S*), climate (*C*) and their interactions (*S* × *C*) in early growth rate were estimated by using adjusted R-squared in redundancy analysis ordination. The goodnesss of fitting in SEM at China’s province level was that *Chi-square* = 23.50, *P* = 0.13, *CFI* = 0.90 and *RMSEA* = 0.13, while state level was that *Chi-square* = 22.65, *P* = 0.12, *CFI* = 0.94 and *RMSEA* = 0.13. These variables were shared by the province and state level, including both climate and socio-economic factors. For climate variables, there were monthly average highest temperature (Avg.HT), monthly average precipitation (Avg.Pre). For socio-economic,there were GDP per capita (GDP.pc), GDP, population (Pop), population density (Pop.density) and commuting rate (Commuting). † The fixed indicators of latent variables. \*\*\**P* < 0.01, ** *P* < 0.05.

Overall, climate and socio-economic factors jointly explained more spatial variations in early growth rate for China and the other counties, that is 54.3% and 32.8%, respectively (Fig. 1b). It means that the predictive certainty of COVID-19 trainsmissions under strict interventions in China was 1.6 times higher than the other countries with less interventions. Such a finding shows that the aspects of containment measures, public health policies and immigration monitoring may inhibit the local transmission dynamics of COVID-19 [2], which are consistent with the findings in the spread of influenza [10] and Zika [9]. In addition, the low explained variations in early growth rate for the other countries may be caused by high mobility which makes local transimission dynamics to be homogeneous [32].

To further test the effects of strict quarantine on the spatial variations of COVID-19 growth rate, we implemented the Global Epidemic and Mobility Model (GLEAM), an stochastic and individual-based model using a metapopulation network to simulate epidemic spread [15, 33, 34]. In the metapopulation networks, nodes or subpopulations were connected by traveller fluxes, and characterized by random socio-economic and climate factors which shape the exponential growth rate of COVID-19 (Fig. 2a). As expected, GLEAM results show that the explained variations of COVID-19 growth rate decreased towards the elevated quarantine level (ANOVA, *P* < 0.001; Fig 2b). This is consistent with empirical results and indicates strict quarantine to enhance the predictability of COVID-19 transmission. In general, high mobility would disturb the heterogeneous structure of metapopulation network [35], cause the increase in synchronization among subpopulations due to the strong spatial coupling in epidemic transmssions, like the spread of influenza in European [36], and thus decrease the predictability in epidemic transmissions [37, 38].

**Figure 2.**
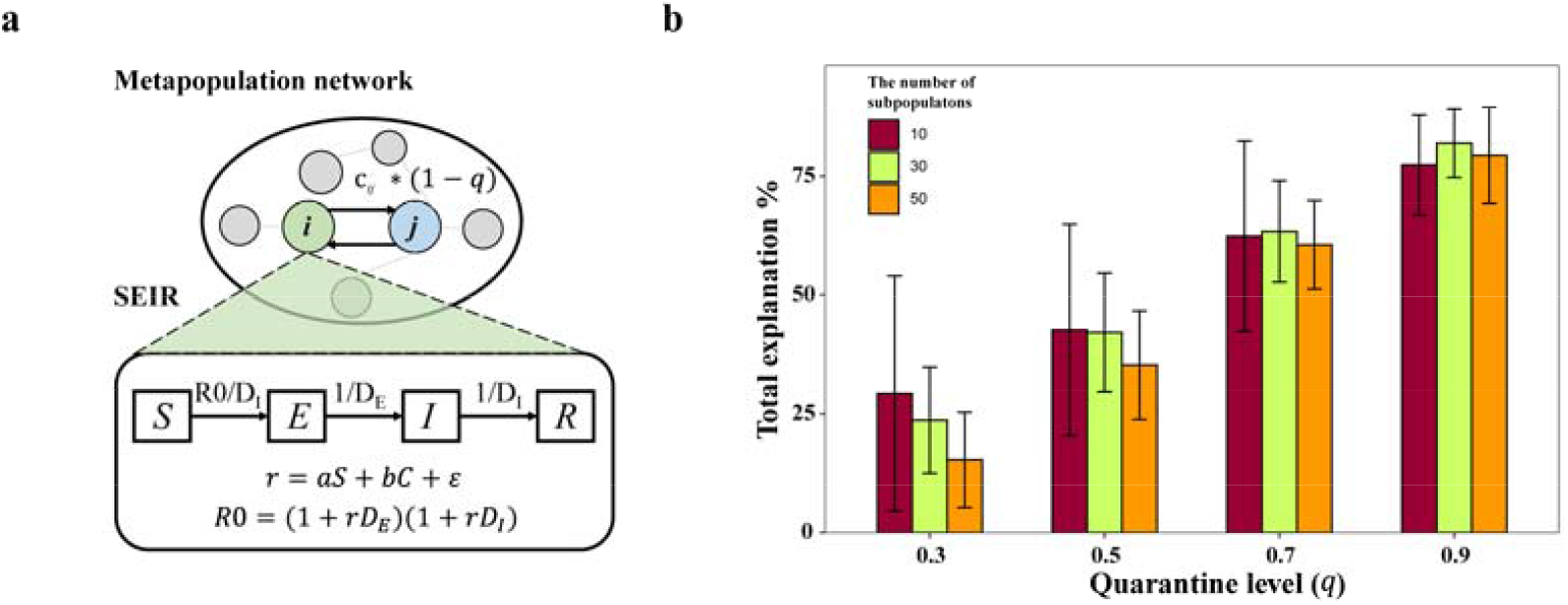
The effects of quarantine regarding travel restrictions on the explainable variations in the early growth rates of COVID-19 driven by socio-economic (S) and climate (C) factors. (**a**) In metapopulation network model, the proportion of immigrants in subpopulation *i* from *j* was *c*_*ji*_ constrained by quarantine level *q*. For each subpopulation, COVID-19 spread was modeled by susceptible-exposed-infected-removed (SEIR) model, where DE and DI were the exposed and infected periods of COVID-19, respectively. R0 is basic reproductive number, and is shaped by random socio-economic (*S*) and climate (*C*) factors with the constants of *a* and *b* and random error *ε*. (**b**) The total variations in early growth rate explained by socio-economic and climate factors with different quarantine level. The quarantine level of zero indicates that there were no travel restrictions among subpopulations, while the quarantine level of one indicate that travel flows among subpopulations are zero. In simulation process, the travel restrictions would be implemented after 20 days of epidemic outbreak and the growth rate were estimated based on accumulative cases during the period of travel restriction. The number of subpopulations in a metapopulation are denoted by the colors of bars.

Although we here show a clear picture on how COVID-19 growth rate was shaped by climate and socio-economic factors and constrained by strict quarantine, two major caveats could be acknowledged regarding the analytical framework. First, COVID-19 growth rate could be estimated by various methods, such as exponential [4, 17, 27] or logistic model [39, 40], and basic reproductive number *R0* [17, 26, 41]. Here, we chose exponential model with extensible windows and determined COVID-19 growth rate as maximum growth rate in logistic growth curve during the exponential phase. In this phase, COVID-19 spread was majorly dependent on intrinsic growth rate of virus, regardless of susceptible population resources associated with external control measures, such as quarantine and vaccination, and thus the maximum growth rate is more accurate to explore the nature potential of virus. We did not consider logistic model as it failed in estimating COVID-19 growth rate during early epidemic. For other models, such as exponential model with moving windows and *R0* estimated by maximum likelihood method, we obtained similar conlusions as follows. (i) Socio-economic factors played more important role in COVID-19 spread than climate (Fig. S8) and (ii) the variations in early growth rate were explained more in China than the other countries (Fig. S8c).

Second, the growth rate may be affected by the data source of reported cases. For example, not all infected indivuals could be tested due to the insufficient capacity for testing and tracing or asymptomatic cases, which would cause the reported cases less than the number of infected individuals. Further, the imported cases count a considerable proportion of reported cases at the early stage, which may disturb the characteristics of local transmission dynamics in COVID-19 spread. Besides, the protocols for reporting COVID-19 cases may vary across countries, which may result in unmeasured spatial variations resulted from data collection, processing and summary.

## Methods and materials

### Data sources

We analyzed the global time series data on infections from January 22th, 2020 to March 26th, 2020 in Ding Xiang Yuan [42]. We only used the regions with at least one reported case of COVID-19 and five data points to remain in the time series after truncation in order to proceed with model fitting, which limited our analysis to 155 regions (that is, 121 countries and 34 China’s provinces). We further collected 16 and 30 environmental and social-economic variables, respectively, as explanatory factors for COVID-19 spread (Table S1). Climate data at country-level and China’s province-level, such as the monthly average, highest and lowest temperatures, and the monthly average precipitation, were collected from Weatherbase [43]. The variables related to area, elevation, population size and GDP of China’s provinces were available from China’s National Bureau of Statistics [44]. For the other countries, the social-economic variables such as population, GDP, body mass index (BMI), air quality and and climate variables of UV level, were collected from World Health Organization [45]. We estimated the commuting rate of China’s provinces based human migration follows from January 1st, 2020 to January 23th, 2020 available from Github repository of https://github.com/2020-nCoV/RawData. The commuting rates of other countries were eastimated by open-source data of air traveller flow in February 2010, and are available from https://sites.google.com/site/liangmaoufl/data-portal.

### Statistical inferences in the early growth rate of COVID-19

During early epidemic of COVID-19, the cumulative cases are usually characterized by exponential growth (refs). Thus, here, the early growth rate *r* of COVID-19 in each region was estimated by the slope of *ln(cases) = ln(a)* + *rt* line via ordinary least squares linear regression, where *cases* is the cumulative reproted cases of COVID-19 at time *t* and *a* is a constant. We defined the extensible windows as the various durations (with a minimum value of five) of time series data starting from the day of the first record, calculated a seires of growth rate *r* for each window and determined the growth rate of COVID-19 as the maximum and statistically significant value (*P* < 0.05). To identify the robustness of estimations on our conclusions, we also determined the growth rates with another four approaches as follows. (1) In the exponential model with extensible windows, we also determine the growth rate as the second, third and fourth maximum *r values* and the average of top four maximum values. (2) We fitted the exponential model with moving windows that have a fixed size ranged from 7 to 14 and determined early growth rate as the maximum estimation for each moving window. (3) The basic reproduction number *R*0 was calculated by maximum likelihood estimation [46]. The serial interval of COVID-19 in each region was equal to that in Wuhan, China, with a mean of 7.5 days and a standard deviation of 3.4 days [47] and the maximum reproduction number was assumed to be eight. (4) we also fitted the logistic model with extensible and moving windows, but it was failed to estimated early growth rate according to *P* value and determinant coefficients *R*^2^, and thus it was not further considered.

### Quanrantine effects on the predictability in the early epidemic growth rates

To examine the effects of travel restrictions on the predictability in empidemic growth rate, we stochastically simulated the COVID-19 sreapd with the Global Epidemic and Mobility Model (GLEAM). GLEAM model is an individual-based, stochastic, and spatial epidemic model [15, 33]. Metapopulation networks of GLEAM were aggregated by *N* subpopulations, in which there is 100,000 susceptible people and the specifically exponential growth rate *r* driven by local climate (*C*) and socio-economic (*S*) factor, as followed:

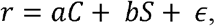

where *a* and *b* are constant and *ϵ* is random error generated by the normal distribution of *N*(0, 0.01). Both of climate and socio-economic factors were generated by the uniform distribution of *U*(0.5, 1). Subpopulations were connected by the traveller fluxes. The flux rate *c*_*ij*_, that is travellers from subpopulation *i* to subpopulation *j* account for the population size of subpopulation *i*, was generated from the logarithmic normal distribution of *LN(*−5, 1).

Human-to-human transmission of epidemic within a subpopulation was modeled by SEIR model that include four compartments including susceptible, latent, infectious and recovered [15]. Individuals in susceptible compartments can acquire the virus through contacts with individuals in infectious compartment and were transfered into latent compartment with a rate *R*_0_*/D*_*I*_, where *R*_0_ is basic reproduction number and *D*_*I*_ is mean infectious period. Latent individuals progress to the infectious stage with a rate *1/D*_*E*_, where *D*_*E*_ is mean latent period, and infectious individuals progress into the recovery stage with a rate 1/*D*_*I*_. Notably, *R*_0_ was obtained through the growth rate *r* with the function of *R*_0_ *=* (1 + *rD*_*E*_)(1 + *rD*_*I*_) [48]. According to previous study [21], the latent period *D*_*E*_ was assumed to be 6.1 days, while infectious period was 1.5 days. Model simulations were performed in Python 3.6.3 [49], while analyses were in R 3.6.1 [50]. Code files are available on https://github.com/zhenghualiu/COVID-19/tree/master/metapopulation_model.

### Statistical analyses

We considered 21 China’s provinces and 26 countries outside of China for downstream analyses. The reasons for selecting these 26 countries are as followed. (i) In these regions, the exponential growth periods of COVID-19 outbreaks were in March and the growth rate could be characterized by average temperature and precipitation of that month. (ii) These countries show the similar area size ranges to the provinces of China (Fig. S5) and could minimize the spatial scale effects on growth rates. (iii) The shared socio-economic variables among these countiers were obtained thus ensuring data integrity. (iv) The similar number of sample points between China and the other countries could reduce the sample size heterogeneity in statistical analyses for solid comparation.

The relationships between COVID-19 growth rate and environmental or socio-economic factors were examined by Pearson’s corelations. To select appropriate predictors from 46 potential variables for determining a minimal adequate model, we filtered the independent variables with high collinearity by showing the absolute values of Pearson’s correlation greater than 0.8. By using random forest model with 500 classification trees, we defined the unimportant variable as a predictor with a relative importance less than 0.5% and excluded the most unimportant variable from exploratory variables. This process was repeated until there are no unimportant variables. Then, the random forest model [51] with the final set of exploratory variables were used to estimate the relative importance of variables to COVID-19 growth rate based on 500 classification trees with 10 cross validations.

The variation in early growth rate explained by variables were estimated by redundancy analysis with the hierarchy algorithm of Chevan and Sutherland [52, 53]. To evaluate the influences of climate and socio-economic factors on COVID-19 spread, the variance in COVID-19 growth rate was further partitioned by two explainatory matrices composed of cimate and socio-economic factors with distance-based redundancy analysis [54]. Further, we constructed the structural equation model (SEM) with maximum-likelihood estimation procedures [55] to quantify the influences of climate and socio-economic latent variables on COVID-19 growth rate. The exploratory variables were shared by both province and state level, including monthly average temperature and precipiatation, GDP per capita, population size, population density and commuting rate. The latent variable of climate was measured by monthly average highest temperature and precipitation, with the loading on monthly average highest temperature constrained to be 1. The socio-economic latent variable was measured by GDP per capita, population, population density and commuting rate, with the latent variable variance fixed to 1 and the loading on GDP per capita constrained to be 1. To acquire minimal adequate model, only the covariances among variables with statistical significance (*P* < 0.1) were kept for SEM fitting (Table S5, S6). The goodnesss of fitting were estimated by *Chi-square* value, the comparative fit index (*CFI*) and the root mean square error of approximation (*RMSEA*).

All above analyses were conducted in R 3.6.1 environment [50]. R codes are available on https://github.com/zhenghualiu/COVID-19/tree/master/Rcodes.

## Data Availability

All data and R codes are available on https://github.com/zhenghualiu/COVID-19/tree/master/Rcodes.

https://github.com/zhenghualiu/COVID-19

## Acknowledgements

I appreciate the contributors for collecting COVID-19 case data around the world, G.N. Wang for helping to collect data and Dr. Liang for providing the data of monthly flows in global air travel passengers.

