## Supplementary material for "Social-economic drivers overwhelm climate in underlying the COVID-19 early growth rate": Table S1, Table S2, Table S3, Table S4

Zhenghua Liu^1,*^

^1^School of Minerals Processing and Bioengineering, Central South University, Changsha 410006, China

*Corresponding authors:

Zhenghua Liu,

**Running title:** Socio-economic and climate factors driving COVID-19 spread

**Supplementary tables and figures**

**Table S1.** The climate and socio-economic factors of China and the other countries outside.

| Abbreviation | Full name of explanatory variable | China | Other countries |
| --- | --- | --- | --- |
| Lat | Latitude | TRUE | TRUE |
| Elev | Elevation (m) | TRUE | TRUE |
| Avg.Tem.Jan | Monthly average temperature in Jan. (℃) | TRUE | TRUE |
| Avg.Tem.Feb | Monthly average temperature in Feb. (℃) | TRUE | TRUE |
| Avg.Tem.Mar | Monthly average temperature in Mar. (℃) | TRUE | TRUE |
| Avg.Pre.Jan | Monthly average precipitation in Jan. (mm) | TRUE | TRUE |
| Avg.Pre.Feb | Monthly average precipitation in Feb. (mm) | TRUE | TRUE |
| Avg.Pre.Mar | Monthly average precipitation in Mar. (mm) | TRUE | TRUE |
| Avg.HT.Jan | Monthly average high temperature in Jan. (℃) | TRUE | TRUE |
| Avg.HT.Feb | Monthly average high temperature in Feb. (℃) | TRUE | TRUE |
| Avg.HT.Mar | Monthly average high temperature in Mar. (℃) | TRUE | TRUE |
| Avg.LT.Jan | Monthly average low temperature in Jan. (℃) | TRUE | TRUE |
| Avg.LT.Feb | Monthly average low temperature in Feb. (℃) | TRUE | TRUE |
| Avg.LT.Mar | Monthly average low temperature in Mar. (℃) | TRUE | TRUE |
| UV | Ultraviolet radiation levels |  | TRUE |
| Extreme.Climate | Droughts, floods, extreme temps (% pop. avg. 1990-2009) |  | TRUE |
| GDP.pc | GDP per capita ($) | TRUE | TRUE |
| GDP | GDP ($) | TRUE | TRUE |
| log.GDPpc | log GDP per capita | TRUE | TRUE |
| Pop | Population | TRUE | TRUE |
| Pop.density | Population density (km2 per 10,000 people) | TRUE | TRUE |
| Pop.below5m | Population below 5m (% of total) |  | TRUE |
| Urban,pop | The proportion of urban population (% of total) |  | TRUE |
| PM2.5 | The content of PM10 (ug/m3) | TRUE | TRUE |
| PM10 | The content of PM10 (ug/m3) | TRUE | TRUE |
| All.BMI | Average BMI of both sexes; 18+ years |  | TRUE |
| Female.BMI | Average BMI of female; 18+ years |  | TRUE |
| Male.BMI | Average BMI of male; 18+ years |  | TRUE |
| All.Obesity | Prevalence of obesity among both sexes; 30 (age-standardized estimate) (%) |  | TRUE |
| Female.Obesity | Prevalence of obesity among females; 30 (age-standardized estimate) (%) |  | TRUE |
| Male.Obesity | Prevalence of obesity among males; 30 (age-standardized estimate) (%) |  | TRUE |
| All.Underweight | Prevalence of underweight among both sexes; 30 (age-standardized estimate) (%) |  | TRUE |
| Female.Underweight | Prevalence of underweight among females; 30 (age-standardized estimate) (%) |  | TRUE |
| Male.Underweight | Prevalence of underweight among males; 30 (age-standardized estimate) (%) |  | TRUE |
| Impro.sanit | Access to improved sanitation (% of total pop.) |  | TRUE |
| Impro.water | Access to improved water source (% of total pop.) |  | TRUE |
| Energy.pc | Energy use per capita (kilograms of oil equivalent) |  | TRUE |
| Energy.pGDP | Energy use per units of GDP (kg oil eq./$1,000 of 2005 PPP $) |  | TRUE |
| FDI | Foreign direct investment, net inflows (% of GDP) |  | TRUE |
| Paved.roads | Paved roads (% of total roads) |  | TRUE |
| Physicians | (per 1,000 people) |  | TRUE |
| CO2.pc | CO2 emissions per capita (metric tons) |  | TRUE |
| N2O.pc | N2O emissions per capita |  | TRUE |
| CH4.pc | CH4 emissions per capita |  | TRUE |
| Commuting | Importing population (% of total pop.) | TRUE | TRUE |
| Mortality.under5 | Under-five mortality rate (per 1,000) |  | TRUE |

**Table S2.** The basic reproductive number (R0) of COVID-19 in China and the other countries collected from references.

| First author | Year | Country | R0 | LCL | UCL |
| --- | --- | --- | --- | --- | --- |
| Joseph T Wu et al | 2020 | China^a^ | 2.68 | 2.47 | 2.86 |
| Mingwang Shen et al | 2020 | China^a^ | 6.49 | 6.31 | 6.66 |
| Tao Liu et al | 2020 | China^a^ | 2.9 | 2.32 | 3.63 |
| Tao Liu et al | 2020 | China^a^ | 2.92 | 2.28 | 3.67 |
| Jonathan M. Read et al | 2020 | China^a^ | 3.11 | 2.39 | 4.13 |
| Maimuna Majumder et al | 2020 | China^a^ | 2.55 | 2 | 3.1 |
| WHO | 2020 | China^a^ | 1.95 | 1.4 | 2.5 |
| Shi Zhao et al | 2020 | China^a^ | 2.24 | 1.96 | 2.55 |
| Shi Zhao et al | 2020 | China^a^ | 3.58 | 2.89 | 4.39 |
| Natsuko Imai | 2020 | China^a^ | 2.5 | 1.5 | 3.5 |
| Julien Riou and Christian L. Althaus | 2020 | China^a^ | 2.2 | 1.4 | 3.8 |
| Tang, Biao et al | 2020 | China^a^ | 6.47 | 5.71 | 7.23 |
| Qun Li et al | 2020 | China^a^ | 2.2 | 1.4 | 3.9 |
| Sheng Zhang et al | 2020 | China^a^ | 2.28 | 2.06 | 2.52 |
| Mingwang Shen et al | 2020 | China^a^ | 4.71 | 4.5 | 4.92 |
| Zhanwei Du et al | 2020 | China^a^ | 1.9 | 1.47 | 2.59 |
| Kamalich Muniz-Rodriguez et al | 2020 | China^a^ | 3.3 | 3.1 | 4.2 |
| Can Zhou | 2020 | China^a^ | 2.12 | 2.04 | 2.18 |
| Tao Liu | 2020 | China^a^ | 4.5 | 4.4 | 4.6 |
| Tao Liu | 2020 | China^a^ | 4.4 | 4.3 | 4.6 |
| Ruiyun Li et al | 2020 | China^a^ | 2.23 | 1.77 | 3 |
| Sang Woo Park et al | 2020 | China^a^ | 3.1 | 2.1 | 5.7 |
| Nian Shao et al | 2020 | China^a^ | 3.32 | 3.25 | 3.4 |
| Huijuan Zhou et al | 2020 | China^a^ | 5.5 | 5.3 | 5.8 |
| Alessia Lai et al | 2020 | China^a^ | 2.6 | 2.1 | 5.1 |
| Sung-mok Jung et al | 2020 | China^a^ | 2.1 | 2 | 2.2 |
| Sung-mok Jung et al | 2020 | China^a^ | 3.2 | 2.7 | 3.7 |
| Steven Sanche et al | 2020 | China^a^ | 6.3 | 3.3 | 11.3 |
| Steven Sanche et al | 2020 | China^a^ | 4.7 | 2.8 | 7.6 |
| Albertine Weber et al [1] | 2020 | South Korea | 1.12 | 1.08 | 1.16 |
| Albertine Weber et al [1] | 2020 | Brunei | 6.45 | 4.6 | 8.3 |
| Albertine Weber et al [1] | 2020 | Sweden | 2.85 | 2.2 | 3.5 |
| Aayah Hammoumi et al [2] | 2020 | Morocco | 2.99 | 2.67 | 3.15 |
| Junko Kurita et al [3] | 2020 | Janpan | 2.16 | 1.97 | 2.2 |
| Xiang Zhou et al [4] | 2020 | Italy | 3.09 | - | - |
| Xiang Zhou et al [4] | 2020 | Iran | 3.47 | - | - |
| Xiang Zhou et al [4] | 2020 | South Korea | 3.88 | - | - |
| Xiang Zhou et al [4] | 2020 | Germany | 3.27 | - | - |
| Xiang Zhou et al [4] | 2020 | France | 3.47 | - | - |
| Xiang Zhou et al [4] | 2020 | US | 3.86 | - | - |
| Xiang Zhou et al [4] | 2020 | Spain | 1.69 | - | - |
| Jing Yuan et al [5] | 2020 | Italy | 3.27 | 3.17 | 3.38 |
| Jing Yuan et al [5] | 2020 | Italy | 2.3 | 2.25 | 2.35 |
| Jing Yuan et al [5] | 2020 | France | 3.6 | 3.37 | 3.85 |
| Jing Yuan et al [5] | 2020 | Germany | 3.5 | 3.28 | 3.74 |
| Jing Yuan et al [5] | 2020 | Spain | 3.11 | 2.86 | 3.37 |
| Francisco H. C. Felix et al [6] | 2020 | Brazil | 1.96 | 1.55 | 2.86 |
| YasetCaicedo-Ochoa et al [7] | 2020 | Spain | 2.9 | 2.67 | 3.14 |
| YasetCaicedo-Ochoa et al [7] | 2020 | Italy | 2.83 | 2.7 | 2.96 |
| YasetCaicedo-Ochoa et al [7] | 2020 | Ecuador | 3.95 | 3.7 | 4.13 |
| YasetCaicedo-Ochoa et al [7] | 2020 | Panama | 3.67 | 3.25 | 4.13 |
| YasetCaicedo-Ochoa et al [7] | 2020 | Brazil | 2.91 | 2.6 | 3.23 |
| YasetCaicedo-Ochoa et al [7] | 2020 | Chile | 2.67 | 2.45 | 2.89 |
| YasetCaicedo-Ochoa et al [7] | 2020 | Colombia | 2.67 | 2.38 | 2.98 |
| YasetCaicedo-Ochoa et al [7] | 2020 | Peru | 2.36 | 2.11 | 2.63 |
| YasetCaicedo-Ochoa et al [7] | 2020 | Mexico | 2.42 | 2.14 | 2.72 |
| Mohamed HAMIDOUCHE et al [8] | 2020 | Algeria | 3.28 | 2.28 | 4.27 |
| Edward De Brouwer et al [9] | 2020 | Italy | 2.68 | 2.42 | 3.08 |
| Edward De Brouwer et al [9] | 2020 | Belgium | 2.1 | 1.94 | 2.12 |
| Tridip Sardar et al [10] | 2020 | India | 2:25 | 1.97 | 4.72 |
| Sunhwa Choi et al [11] | 2020 | South Korea | 0.56 | 0.51 | 0.60 |
| Eunha Shim et al [12] | 2020 | South Korea | 1.5 | 1.4 | 1.6 |
| Jia Wangping et al [13] | 2020 | Italy | 4.34 | 3.04 | 6.00 |
| Samuel P C Brand et al [14] | 2020 | Kenya | 3.46 | 2.81 | 4.17 |
| Durgesh Nandini Sinha [15] | 2020 | World | 2.56 | 2.12 | 2.99 |

^a^ The R0 in China were collected by Yousef Alimohamadi et al [16].

**Table S3.** The correlations between the COVID-19 early growth rate and socio-economic or climate factors in the countries except for China.

| Variables | Pearson's r | p.value |
| --- | --- | --- |
| **Lat** | **0.38** | **0.04** |
| Elev | 0.00 | 0.99 |
| Avg.Tem.Jan | -0.34 | 0.07 |
| Avg.Tem.Feb | -0.34 | 0.07 |
| **Avg.Tem.Mar** | **-0.37** | **0.05** |
| Avg.Pre.Jan | 0.17 | 0.38 |
| Avg.Pre.Feb | 0.22 | 0.26 |
| Avg.Pre.Mar | 0.17 | 0.38 |
| Avg.HT.Jan | -0.36 | 0.06 |
| Avg.HT.Feb | -0.36 | 0.06 |
| **Avg.HT.Mar** | **-0.39** | **0.04** |
| Avg.LT.Jan | -0.32 | 0.09 |
| Avg.LT.Feb | -0.32 | 0.10 |
| Avg.LT.Mar | -0.35 | 0.06 |
| **UV** | **-0.42** | **0.02** |
| Extreme.Climate | -0.35 | 0.07 |
| **GDP.pc** | **0.63** | **0.00** |
| GDP | -0.18 | 0.35 |
| Pop | **-0.37** | **0.05** |
| Pop.density | -0.07 | 0.72 |
| Urban.pop | 0.31 | 0.10 |
| All.BMI | 0.25 | 0.20 |
| Female.BMI | 0.12 | 0.54 |
| Male.BMI | 0.35 | 0.07 |
| All.Obesity | 0.25 | 0.19 |
| Female.Obesity | 0.16 | 0.42 |
| Male.Obesity | 0.33 | 0.08 |
| All.Underweight | -0.33 | 0.08 |
| Female.Underweight | -0.32 | 0.09 |
| Male.Underweight | -0.33 | 0.08 |
| Commuting | **0.54** | **0.00** |
| PM2.5 | -0.14 | 0.47 |
| PM10 | -0.11 | 0.57 |
| Impro.sanit | 0.26 | 0.18 |
| Impro.water | 0.14 | 0.47 |
| Energy.pc | **0.51** | **0.00** |
| Energy.pGDP | 0.22 | 0.26 |
| FDI | 0.19 | 0.33 |
| Paved.roads | 0.22 | 0.26 |
| Physicians | 0.35 | 0.06 |
| Mortality.under5 | -0.23 | 0.22 |
| CO2.pc | 0.30 | 0.11 |
| N2O.pc | **0.36** | **0.05** |
| CH4.pc | 0.31 | 0.10 |

**Table S4.** The correlations between the COVID-19 early growth rate and socio-economic or climate factors in China.

| Variables | Pearson's r | p.value |
| --- | --- | --- |
| Lat | -0.25 | 0.27 |
| Elev | -0.19 | 0.42 |
| Avg.Tem.Jan | 0.22 | 0.34 |
| Avg.Tem.Feb | 0.19 | 0.42 |
| Avg.Tem.Mar | 0.14 | 0.54 |
| Avg.Pre.Jan | 0.27 | 0.24 |
| Avg.Pre.Feb | 0.31 | 0.17 |
| Avg.Pre.Mar | 0.31 | 0.18 |
| Avg.HT.Jan | 0.22 | 0.34 |
| Avg.HT.Feb | 0.22 | 0.34 |
| Avg.HT.Mar | 0.16 | 0.50 |
| Avg.LT.Jan | 0.22 | 0.33 |
| Avg.LT.Feb | 0.19 | 0.41 |
| Avg.LT.Mar | 0.15 | 0.51 |
| **GDP.pc** | **-0.52** | **0.01** |
| GDP | -0.02 | 0.95 |
| Commuting | -0.15 | 0.51 |
| Pop | 0.16 | 0.50 |
| Pop.density | -0.37 | 0.10 |
| PM2.5 | 0.02 | 0.93 |
| PM10 | 0.09 | 0.69 |

**Table S5.** The summary of structural equation models for China.

|  | Variables | Estimate | Std.Err* | z-value | P(>\|z\|) | Std.lv* | Std.all* |
| --- | --- | --- | --- | --- | --- | --- | --- |
| Latent Variables: | | | | | | | |
| Climate | Avg.HT.Jan | 1 |  |  |  | 1.358 | 1.392 |
|  | Avg.Pre.Jan | 0.284 | 0.452 | 0.629 | 0.529 | 0.386 | 0.396 |
| Socio-economic | GDP.pc | 1 |  |  |  | 0.89 | 0.912 |
|  | GDP | -0.176 | 0.251 | -0.701 | 0.483 | -0.157 | -0.161 |
|  | Pop.density | 0.184 | 0.256 | 0.718 | 0.473 | 0.164 | 0.168 |
|  | **Commuting** | **0.715** | **0.23** | **3.103** | **0.002** | **0.636** | **0.652** |
|  | **Pop** | **-0.507** | **0.237** | **-2.143** | **0.032** | **-0.451** | **-0.463** |
| Regressions: | | | | | | | |
| Growth rate | Climate | -0.05 | 0.13 | -0.382 | 0.702 | -0.068 | -0.069 |
|  | **Socio-economic** | **-0.606** | **0.245** | **-2.474** | **0.013** | **-0.539** | **-0.552** |
| Covariances: |  |  |  |  |  |  |  |
| **GDP** | **Pop** | **0.683** | **0.239** | **2.86** | **0.004** | **0.683** | **0.82** |
| **Pop.density** | **Commuting** | **-0.555** | **0.203** | **-2.739** | **0.006** | **-0.555** | **-0.78** |
| **Climate** | **Socio-economic** | **-0.385** | **0.22** | **-1.75** | **0.08** | **-0.318** | **-0.318** |
| Variances | | | | | | | |
|  | Socio-economic | 0.792 | 0.316 | 2.503 | 0.012 | 1 | 1 |
|  | Climate | 1.845 | 2.719 | 0.679 | 0.497 | 1 | 1 |
|  | Avg.HT.Jan | -0.893 | 2.731 | -0.327 | 0.744 | -0.893 | -0.938 |
|  | Avg.Pre.Jan | 0.803 | 0.331 | 2.427 | 0.015 | 0.803 | 0.844 |
|  | GDP.pc | 0.16 | 0.136 | 1.175 | 0.24 | 0.16 | 0.168 |
|  | GDP | 0.928 | 0.287 | 3.23 | 0.001 | 0.928 | 0.974 |
|  | Pop.density | 0.926 | 0.287 | 3.225 | 0.001 | 0.926 | 0.972 |
|  | Commuting | 0.547 | 0.194 | 2.818 | 0.005 | 0.547 | 0.575 |
|  | Pop | 0.749 | 0.239 | 3.133 | 0.002 | 0.749 | 0.786 |
|  | Growth rate | 0.681 | 0.221 | 3.075 | 0.002 | 0.681 | 0.715 |

*Std.Err: Standard error; Std.lv: Standard error by latent variable; Std.all: Standard error by all covariances and variances

**Table S6.** The summary of structural equation models for the countries outside of China.

|  | Variables | Estimate | Std.Err* | z-value | P(>\|z\|) | Std.lv* | Std.all* |
| --- | --- | --- | --- | --- | --- | --- | --- |
| Latent Variables: | | | | | | | |
| Climate | Avg.HT.Mar | 1 |  |  |  | 1 | 1.071 |
|  | Avg.Pre.Mar | 0.166 | 0.188 | 0.879 | 0.379 | 0.166 | 0.162 |
| Socio-economic | GDP.pc | 1 |  |  |  | 0.956 | 0.99 |
|  | GDP | 0.056 | 0.196 | 0.288 | 0.773 | 0.054 | 0.064 |
|  | Pop.density | -0.23 | 0.173 | -1.33 | 0.183 | -0.22 | -0.219 |
|  | Commuting | 0.805 | 0.134 | 5.999 | 0 | 0.769 | 0.82 |
|  | Pop | -0.607 | 0.179 | -3.393 | 0.001 | -0.58 | -0.577 |
| Regressions: | | | | | | | |
| Growth rate | Climate | -0.005 | 0.183 | -0.03 | 0.976 | -0.005 | -0.006 |
|  | **Socio-economic** | **0.413** | **0.192** | **2.154** | **0.031** | **0.395** | **0.436** |
| Covariances: | | | | | | | |
| GDP.pc | GDP | 0.258 | 0.106 | 2.427 | 0.015 | 0.258 | 2.293 |
| GDP | Commuting | 0.201 | 0.092 | 2.182 | 0.029 | 0.201 | 0.446 |
| GDP | Pop | 0.354 | 0.167 | 2.116 | 0.034 | 0.354 | 0.514 |
| Climate | Socio-economic | -0.66 | 0.212 | -3.112 | 0.002 | -0.691 | -0.691 |
| Variances: | | | | | | | |
|  | Socio-economic | 0.913 | 0.269 | 3.395 | 0.001 | 1 | 1 |
|  | Climate | 1 |  |  |  | 1 | 1 |
|  | Avg.HT.Mar | -0.128 | 0.233 | -0.552 | 0.581 | -0.128 | -0.147 |
|  | Avg.Pre.Mar | 1.014 | 0.28 | 3.619 | 0 | 1.014 | 0.974 |
|  | GDP.pc | 0.018 | 0.085 | 0.212 | 0.832 | 0.018 | 0.019 |
|  | GDP | 0.705 | 0.178 | 3.961 | 0 | 0.705 | 0.996 |
|  | Pop.density | 0.958 | 0.262 | 3.656 | 0 | 0.958 | 0.952 |
|  | Commuting | 0.288 | 0.097 | 2.96 | 0.003 | 0.288 | 0.327 |
|  | Pop | 0.675 | 0.191 | 3.536 | 0 | 0.675 | 0.667 |
|  | Growth rate | 0.662 | 0.176 | 3.758 | 0 | 0.662 | 0.806 |

*Std.Err: Standard error; Std.lv: Standard error by latent variable; Std.all: Standard error by all covariances and variances

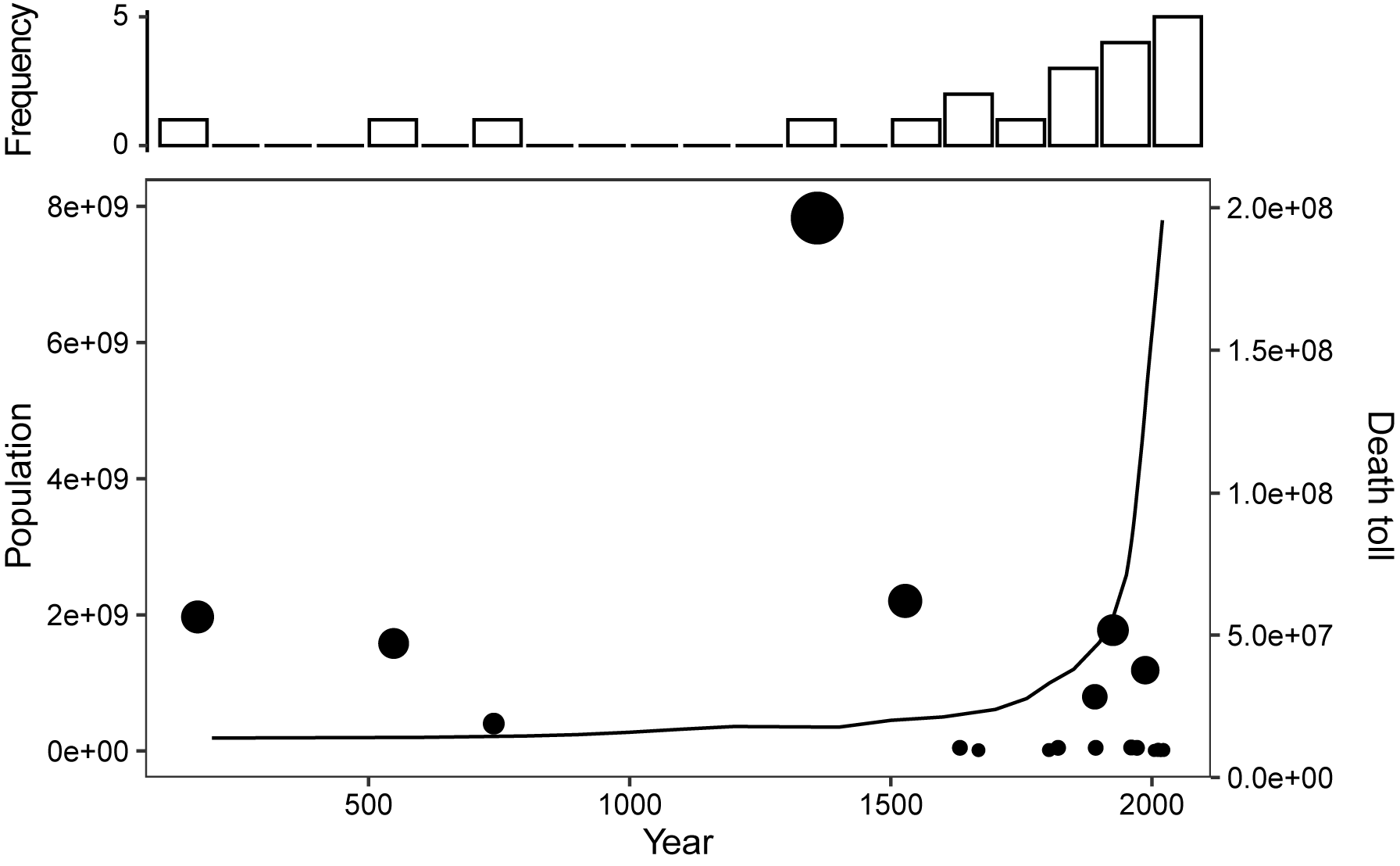

**Figure S1.** The frequency (upper panel) and death tolls (lower panel) of epidemic outbreaks. The Line is population size over time.

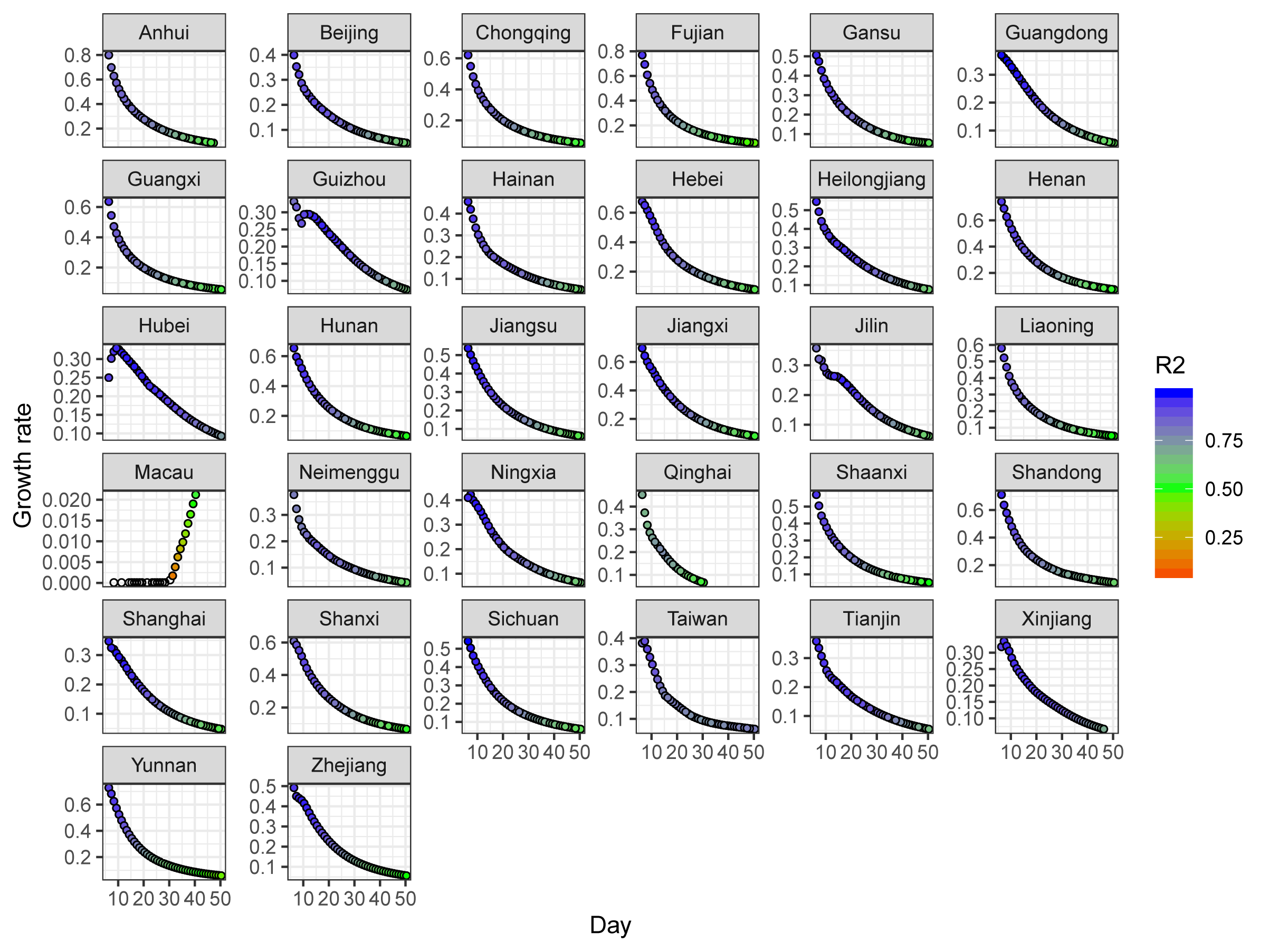
**Figure S2.** The growth rate of COVID-19 estimated by exponential model with extensible window for China. The colors of points represent the changes in R-square, while the white points indicate that the growth rates which were not significant (*P* > 0.05).

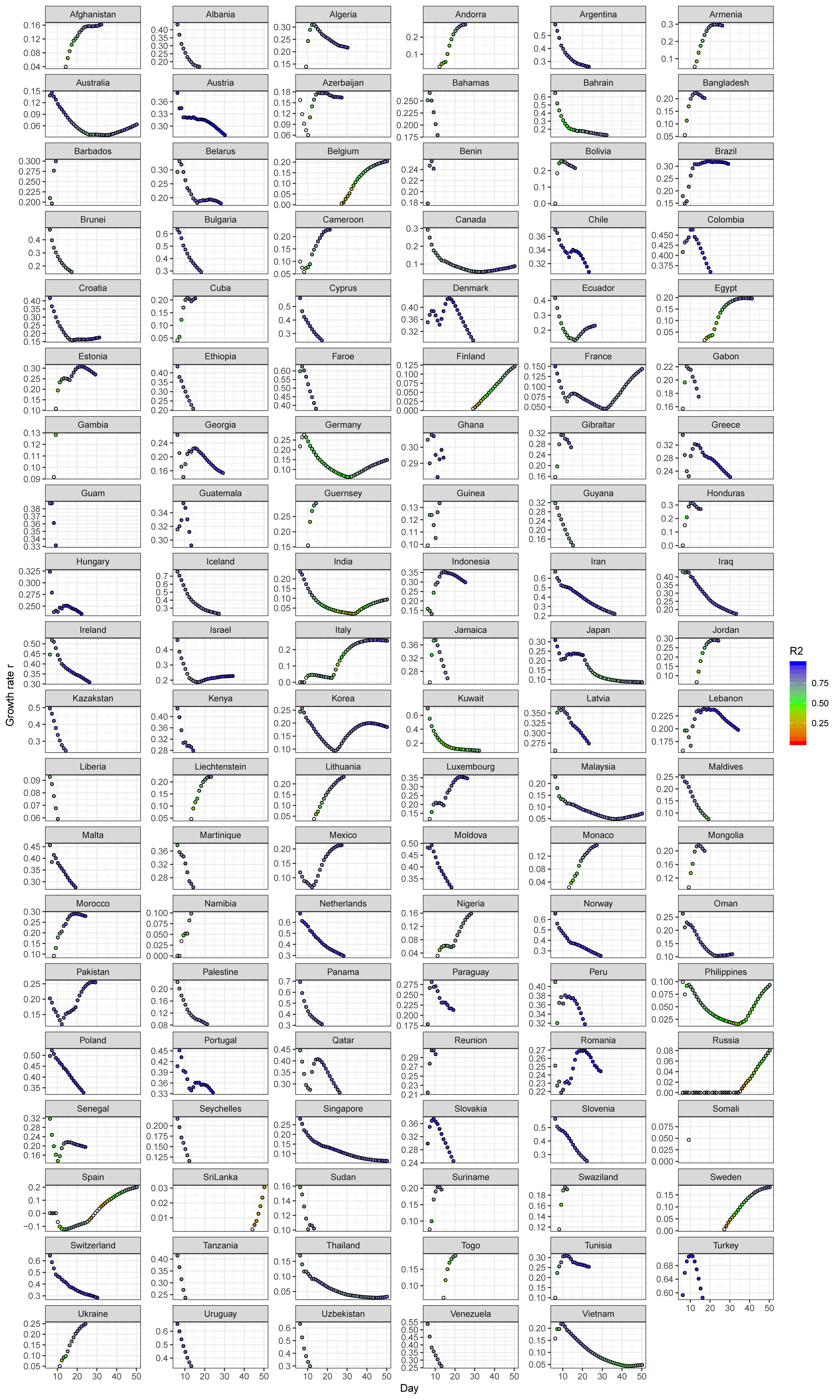

**Figure S3.** The growth rate of COVID-19 estimated by exponential model with extensible window for the countries outside. The colors of points represent the R-square, while the white points denote the growth rate which are not significant (*P* > 0.05).

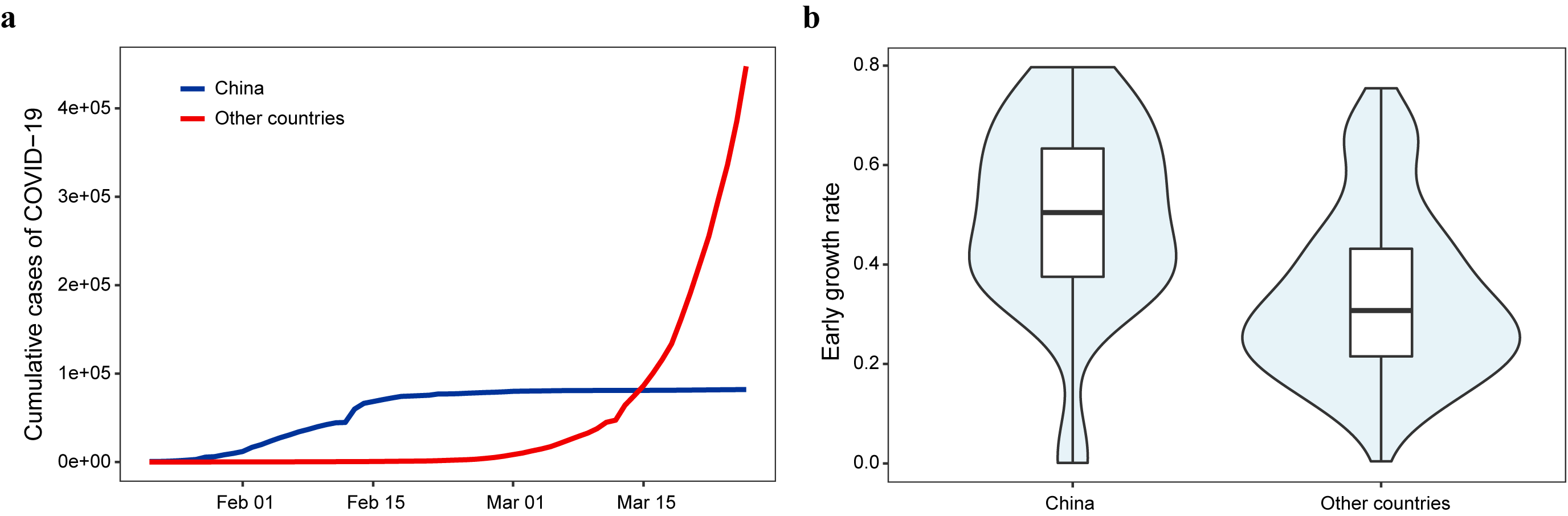
**Figure S4**. The curves of cumulative confirmed infections (**a**) and the box plot of early growth rates (**b**) in China and the other countries.

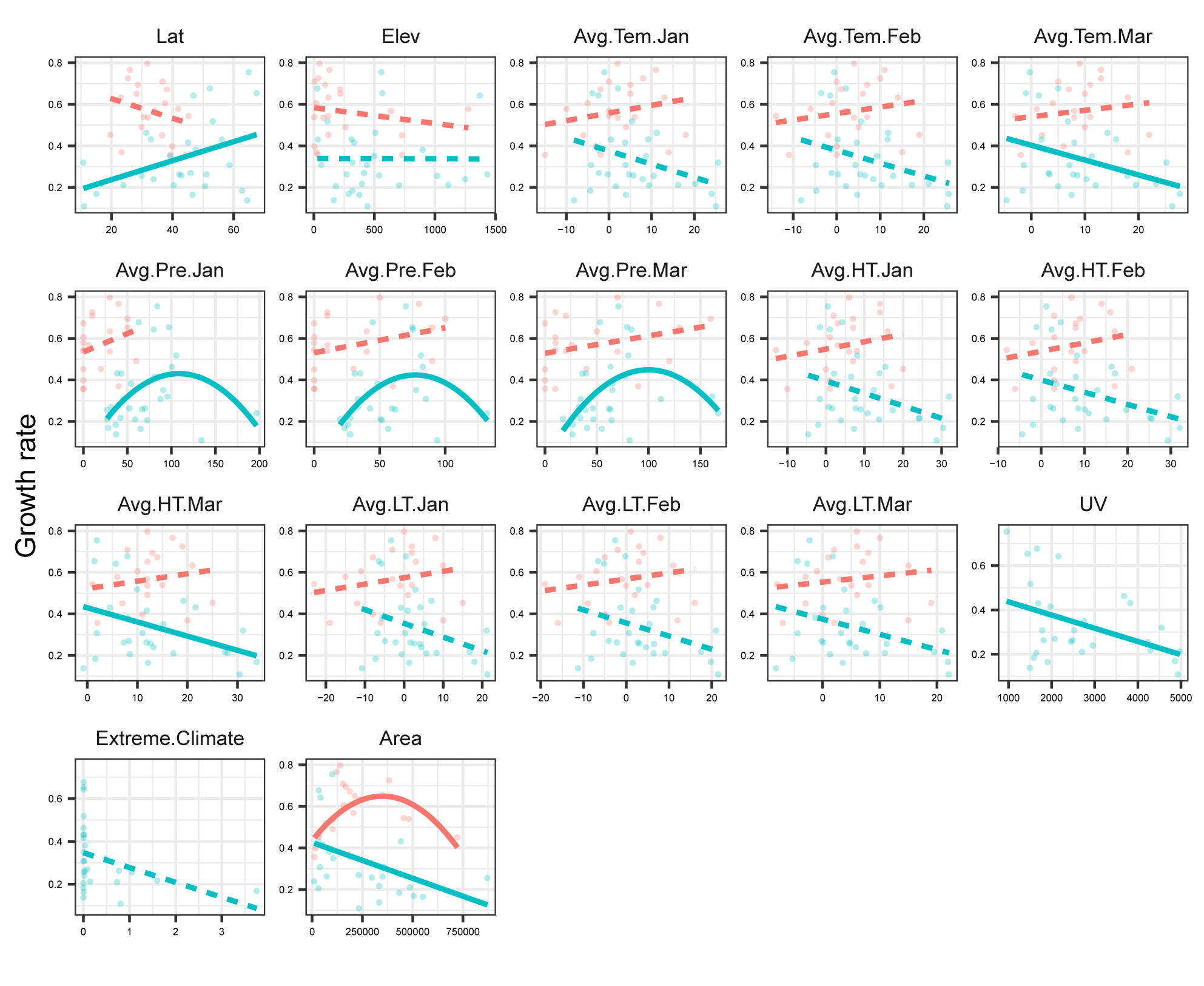
**Figure S5.** The relationships between the COVID-19 early growth rate and climate or environment factors. The red lines or points are the growth rates for the provinces of China, while blue for the other countries. The dashed curves denote that there are no significant (*P* > 0.05) linear or quadratic relationships between the early growth rate and variables, while solid lines indicate significant relationships (*P* < 0.05).

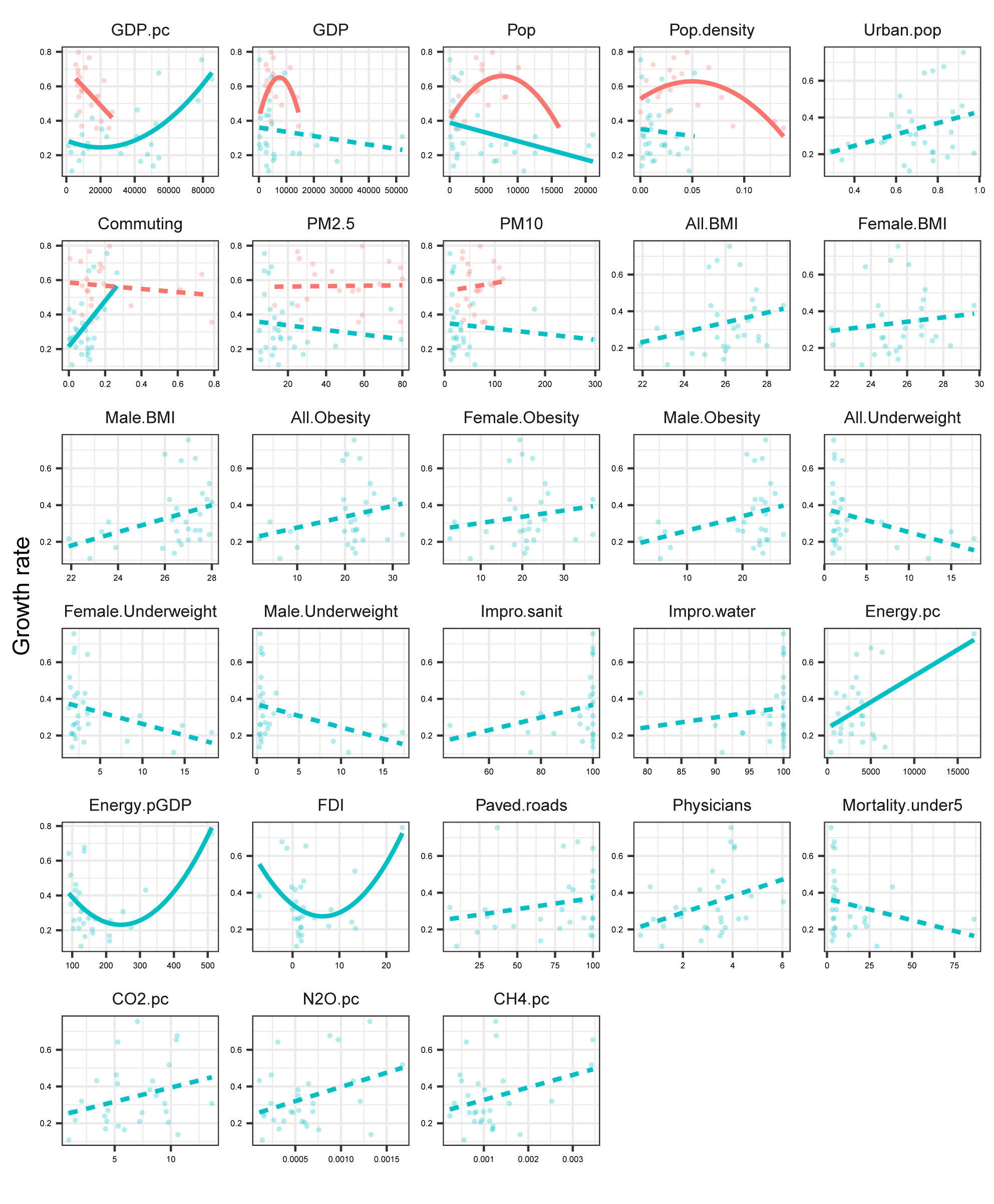
**Figure S6.** The relationships between the COVID-19 early growth rate and socio-economic factors. The red lines or points are the growth rates for the provinces of China, while blue for the other countries. The dashed curves denote that there are no significant (*P* > 0.05) linear or quadratic relationships between the early growth rate and variables, while solid lines indicate significant relationships (*P* < 0.05).

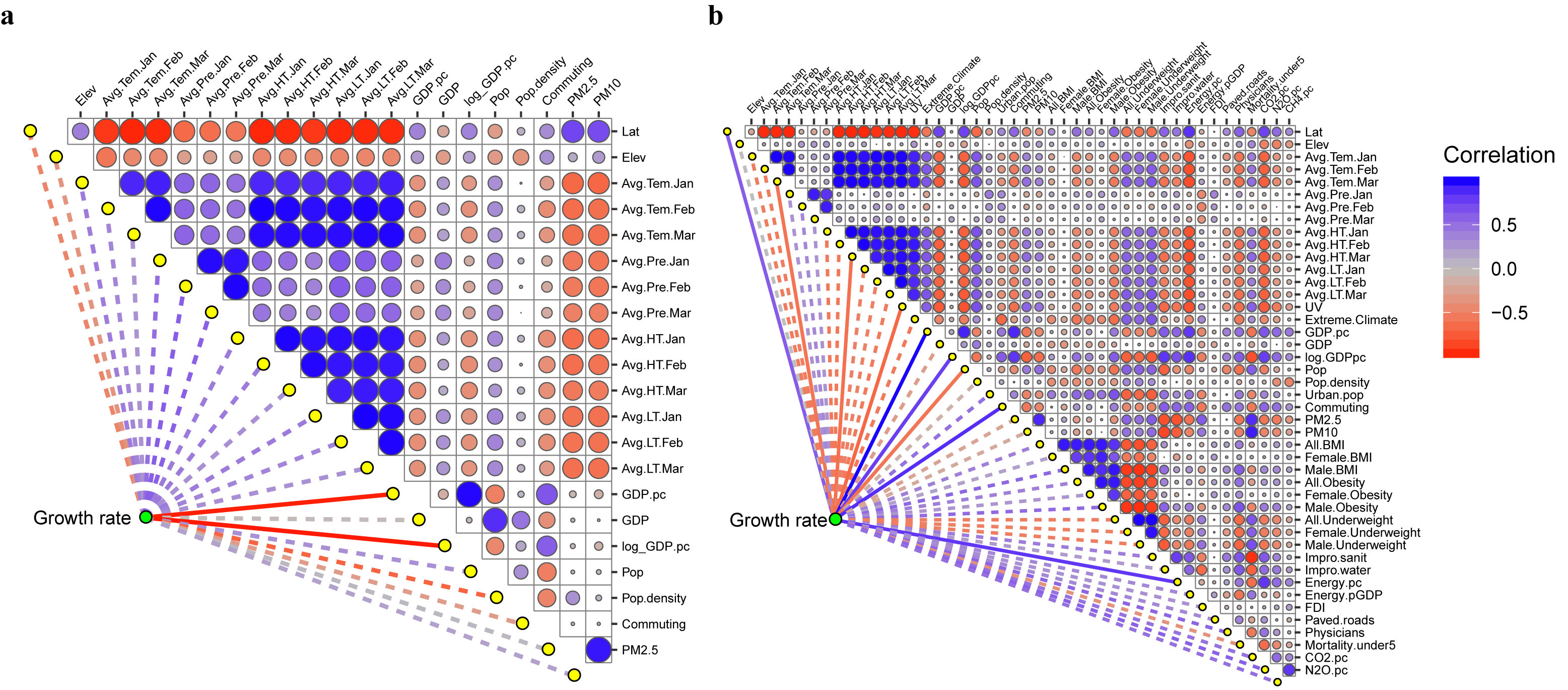

**Figure S7.** The correlations among variables for China (**a**) and the other countries (**b**). The size and color of circles represent the Pearson’s correlations. The solid lines indicate the relationships between early growth rate and explanatory variables estimated by linear regressions which are significant (*P* < 0.05), and dashed lines for non-significant (*P* > 0.05).

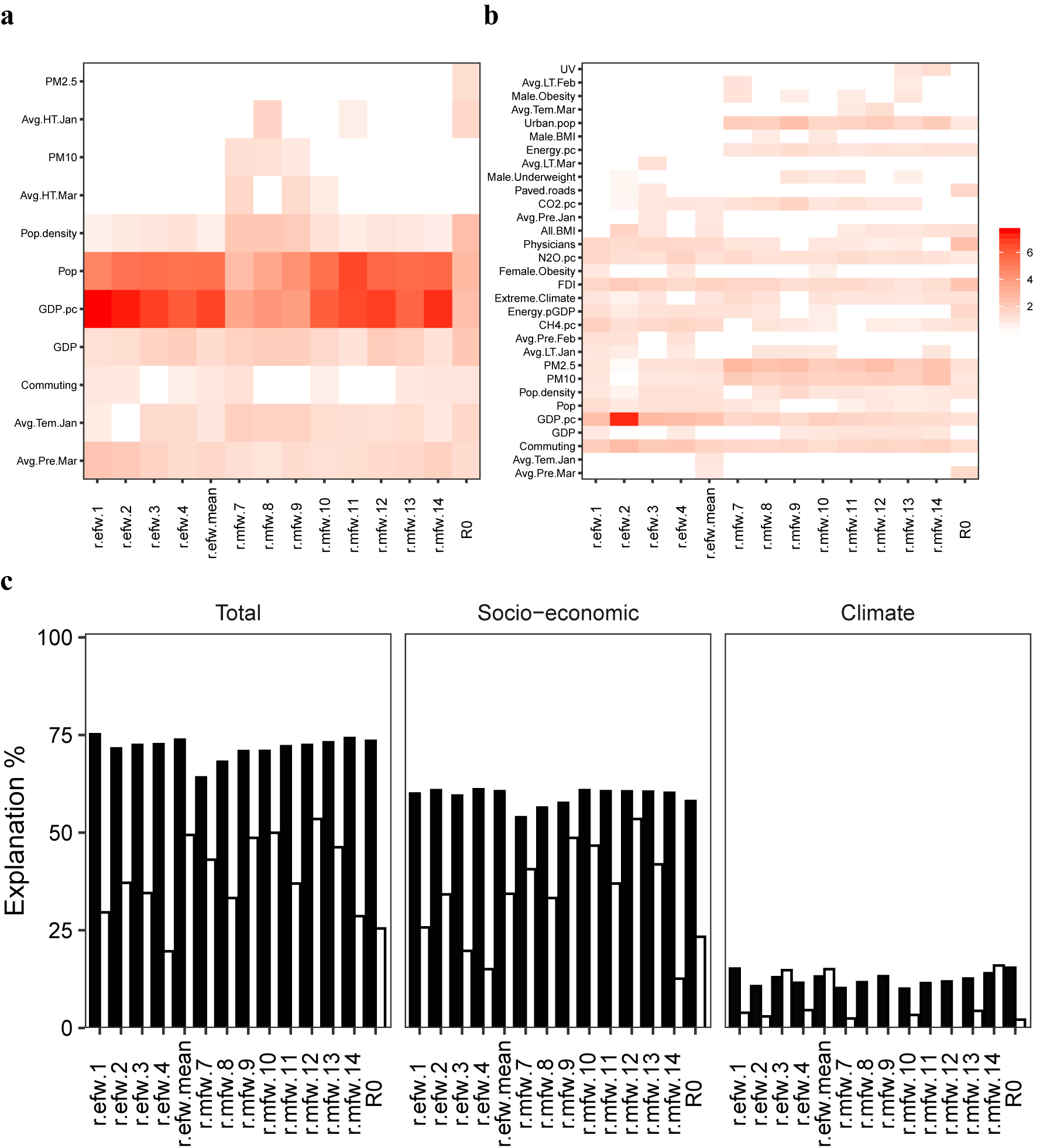
**Figure S8.** The relative importance of variables and total explained variations in COVID-19 growth rate estimated by three methods. These methods are the follows: (1) In the exponential model with extensible windows, the early growth rates, shorten as *r.efw.n*, are determined as the top *n* maximum growth rate, and *n* = 1 denotes the maximum growth rate. The average value of *r.efw.n* is referred as *refw.mean*. (2) In the exponential model with moving windows, the early growth rates, shorten as *r.mfw.m*, are determined as the maximum growth rate for each moving window with size *m*. (3) *R0* is the basic reproduction number estimated by maximum likelihood method.
